# Estimating the death toll of the Covid-19 pandemic in India

**DOI:** 10.1101/2021.06.29.21257965

**Authors:** Christophe Z Guilmoto

**Affiliations:** Centre des Sciences Humaines, Delhi; Ceped/IRD/Université de Paris/INSERM, Paris

## Abstract

The absence of reliable registration of Covid-19 deaths in India has prevented the proper assessment and monitoring of coronavirus pandemic. India’s relatively young age structure tends to conceal the severity of Covid-19 mortality, which is concentrated in older age groups. In this paper, we present four different demographic samples of Indian populations for which we have information on both their demographic structures and death outcomes. We show that we can model the age gradient of Covid-19 mortality in India and use this modeling for estimating the level of Covid-19 mortality in the country. Our findings point to a death toll of about 2.2 million persons by late May 2021. Once India’s age structure is taken into account, these figures correspond to one of the most severe cases of Covid-19 mortality in the world.

**Background:** India has recorded after February a second outbreak of coronavirus that has affected the entire country. The accuracy of official statistics of Covid-19 mortality has been called in question and the real number of Covid-19 deaths is thought to be several times higher than reported. In this paper, we assembled four independent population samples to model and estimate the level of Covid-19 mortality in India.

**Methods:** We first used a first population sample with age and sex of Covid-19 victims to develop a Gompertz model of Covid-19 mortality in India. We applied and adjusted this mortality model on two other national population samples after factoring in the demographic characteristics of these samples. We finally derive from one of these samples the most reasonable estimate of Covid-19 mortality level in India and confirm this result with the use of a fourth population sample.

**Findings:** Our findings point to a death toll of about 2.2 million persons by late May 2021. This is the largest number of Covid-19 deaths in the world. Once standardized for its age and sex structure, India’s Covid-19 mortality rate is above that of Brazil or the USA.

**Interpretation:** Our analysis shows that existing population samples allow for an alternative estimation of deaths due to Covid-19 in India. The results confirm that only one out 7 Covid-19 deaths appear to be registered in India. The estimates point to a very Covid-19 mortality rate, which is even higher after age and sex standardization. The magnitude of the pandemic in India requires immediate attention and calls for a strong response based on a combination of non-pharmaceutical interventions and the scale-up of vaccination to make them accessible to all, with an improved surveillance system to monitor the progression of the pandemic.

---

India has been affected by a new wave of Covid-19 infections and deaths since February 2021. With the emergence of the Delta variant (B.1.617.2),^**1**^ this second wave has been particularly severe and has hit most of the country, starting with the several urban agglomerations and spreading gradually to rural areas. There is, however, a serious gap between the severity of the crisis as reported by local communities and the effect measured by official indicators of contamination and Covid-19 related deaths. The country has officially registered only 311,000 Covid-19 deaths on May 25, 2020. This number corresponds to a rate of 223 per million inhabitants, which is twice lower than the world average.^**2**^Such a low figure contradicts the apparent severity of a crisis that has struck most Indian families across the country, illustrated by dramatic shortages of vaccines, Covid-19 tests, ambulances, access to health personnel, hospital beds, oxygen, ventilators, drugs and finally of coffins, wood, priests and spots for cremation or burials as widely reported in Indian and international media.^**3**^

The purpose of this paper is to provide an alternative estimation method to compute the tally of Covid-19 deaths in India. We use here four independent population samples to reconstruct the impact of mortality. These samples of various sizes and composition are employed to first characterize the age and sex profile of India’s Covid-19 mortality and then to assess its severity by fitting mortality patterns to the number of deaths reported in these samples. This estimation leads to a death tally of about 2.2 million from March 2020 to end of May 2021, a figure about 7 times higher than the official numbers.

The paper starts with a brief presentation of the issues related to data quality and availability in India. The next section describes the alternative data sources on Covid-19 mortality and our estimation strategy. The final sections present our estimation in terms of deaths and India’s position in comparison with the rest of the world. Methodological details are found in three appendices.

## 1. Absence of Reliable Estimates of Covid-19 Deaths in India

The limitations of India’s civil registration are many. Except for a few states or cities with reasonably good statistical sources, deaths are significantly under-registered across the country for a variety of reasons. Death registration was most recently estimated at 77% in 2015, with registration completeness ranging from 32% (e.g., in Bihar) to 100% (e.g., in Kerala) across states.^**4**^ In addition, death statistics are published several years after their occurrence (no national figure is available on the number of deaths in 2019 or 2020) and with minimal information (if any) on the basic characteristics of deaths by age, sex, date, or cause.

In addition to the overall level of under-registration, cases of Covid-19 deaths are known to have been specifically underestimated for two main reasons: many deaths go unrecorded for lack of prior PCR test of the deceased; the cause of death is often selectively attributed to co-morbidities (diabetes, asthma, etc.) and other apparent factors (heart attacks, etc.) when the WHO recommends suspected cases of Covid-19 to be reported as such.^**5 6 7 8 9**^ More generally, Covid-19 remains a source of shame and families often prefer to conceal the nature of the ailment affecting their relatives, ascribing resulting deaths to fevers or other diseases to avoid the social stigma. ^**10 11 12**^ This embarrassment is also felt at political level and Covid-19 figures have been used from the start of the crisis as a political chip to blame national and regional governments for wrong decisions or inaction or even to target minorities.^**13**^ This context provides no incentive at both micro and macro levels to record faithfully and exhaustively cases of Covid-19. They are therefore no instrument to monitor adequately episodes of excess mortality at national level and we may not soon know the true extent of the crisis unless a specific survey is launched this year. ^**14 15**^

During the latest bout of mortality in March-May 2021, the press and civil organizations have actively reported the discrepancies between official figures of Covid-19 mortality and local evidence, resorting to innovative sources such as daily deaths counted in hospitals, number of cremations and burials, corpses found floating in rivers, or obituaries published in local newspapers. While highlighting the gaps in the death counts, this indirect evidence has not led to precise estimates at the aggregate level. ^**16**^

The lack of reliable data has already led to alternative estimates. Available estimates are based on cross-regional modeling and “expert opinion” that are difficult to independently validate for India. ^**17 18 19**^ The only series of estimates based on actual Indian mortality data uses excess mortality measurements in 6 cities (Bengaluru, Chennai, Delhi, Kolkata Mumbai, Nagpur) and one state, leading to a total of 1.8-2.4 million deaths in India by May 16.^**20**^ This estimation procedure remains, however, fragile as it leaves open at least two questions. The first relates to the representativeness of these areas for India as a whole, given that they represent some of the densest and most prosperous urban areas in a country that is 65% rural. The second issue is about the reliability of measurements of Covid-19 deaths based on excess mortality computed as the difference between observed and expected mortality. Non-Covid mortality is most likely to decrease significantly during the pandemic–notably due to lesser exposure to other mortality risks due to lockdown and safety precautions–, which will in turn lead to an underestimation of actual Covid-19 mortality. There are additional issues related the basic measurement of expected mortality based on past trends that may poorly be measured in India by civil registration data. An obvious illustration of these limitations comes from the low mortality experience of Kerala in 2020 and the apparent absence of any excess mortality in this part of India during the pandemic (on Kerala’s Covid-19 mortality, see Appendix 2).

## 2. Using Alternative Sources on Covid-19 Mortality in India

In this study, we identify new sources on Covid-19 mortality to develop reliable estimates of the deaths in India since the beginning of the pandemic. We compute death rates applied to existing population samples for which we have both the Covid-19 death toll (the “numerator”) and the demographic characteristics of the sample (the “denominator”). After correcting for the effect of age and sex structure, we have an estimate of Covid-19 mortality which may be applicable to a specific region or to India as a whole.

Existing sources have carefully been screened for reliability, exhaustivity and representativeness (Appendix 1). To provide a reliable basis for mortality estimation, these samples should meet several criteria:

1. Samples based on well-defined populations (denominator).
2. Reliable estimation of the Covid-19 deaths (numerator).
3. Samples large enough for mortality estimates (robustness).
4. Populations with known age and sex structures (demographic composition)
5. Samples covering the entire pandemic since March 2020 (time scale).
6. Samples representative at all-India level (coverage).

We examined eleven potential sources for estimating Covid-19 mortality, but we had to reject most of them, including those based on deaths among the Indian Armed Forces, health personnel, the schoolteachers of Uttar Pradesh, India’s bank employees, the employees of Hindustan Aeronautics Limited and civil aircraft pilots, and finally the death claims registered by insurance companies. They were mostly rejected to last three criteria listed above (demographic information, time coverage, and representativeness) .

We retained only four different sources.

A. Kerala sample: Dataset of deaths by age and sex in Kerala (May 18; 6,339 deaths)
B. MLA sample: Deaths of elected representatives (May 20; 43 deaths)
C. IR sample: Deaths of Indian Railways personnel (May 7; 1,952 deaths)
D. Karnataka sample: Deaths of schoolteachers in Karnataka (May 13; 268 deaths)

Appendix 1 presents the sources examined and provides a more detailed discussion of the features of these four selected samples. We provide here only a brief description of these samples and of their potential use.

A. The **Kerala sample** is the unique dataset of 6,339 Covid-19 deaths classified by age and sex till May 18 for the southern state of Kerala. It comes from the state with a strong public health system, whose response to the outbreak has been internationally lauded.^**21**^ Demographic data are always more reliable in Kerala than in the rest of India due to its highly educated population. This sample will be used only to determine the age gradient of Covid-19 mortality by modeling death rates.
B. The **MLA sample** (where MLA stands for Member of the Legislative Assembly) consists of 5,837 elected representatives of India. It is composed of members of the regional assemblies (*Vidhan Sabha*) and of the national assemblies (*Lok Sabha* and *Rajya Sabha*). We have both information on individual deaths (by age, sex and date) reported in the press till May 25 as well as the detailed demographic composition of this sample. This sample offers a wide regional representation, but mostly represents a specific elite population. As argued in Appendix 1, we consider this sample to provide an upper bound of Covid-19 mortality in India due to the heavy and prolonged exposure to contamination risks incurred by elected representatives since March 2020.
C. The **IR sample** (where IR stands for Indian Railways) is composed of the 1.25 million permanent workers of the Indian railway sectors and represents the largest sample. Compared to the MLA sample, it provides a far more representative cross-section of India’s population–in terms or regions, social classes, and minorities. It includes a share of frontline workers, but probably no more than the overall Indian workforce. It reported a total number of 1,952 deaths due to the coronavirus till 7 May 2021. Its dedicated health system and potential benefits to families of COVID-19 victims ensure a relatively good level of death registration. This sample will provide the central estimate of Covid-19 mortality in India.
D. The **Karnataka sample** relates to the 268 deaths attributed to Covid-19 among schoolteachers of southern state of Karnataka. These deaths can be related to the demographic characteristics of Karnataka’s 196,000 teachers. Like the Kerala sample, this sample may not reflect the national mortality trends, but will serve as a subsidiary sample to test the validity of mortality estimates.

## 3. Mortality Estimation Strategy

The estimation strategy proceeds from the idea that mortality observed in three samples (MLA, IR and Karnataka) can be simulated after specific demographic adjustments by applying the mortality patterns measured from the Kerala sample. We first apply Kerala’s age- and sex-specific Covid-19 death rates to a sample and then adjust the intensity of mortality to fit the number of observed deaths. The adjusted mortality patterns can finally be applied to India’s age and sex structure to yield national-level estimates of Covid-19 mortality.

This procedure rests on two hypotheses. The first posits that these samples reflect of the severity of Covid-19 mortality in India. As already discussed, we consider the IR the closest to Indian mortality experience because of its social and regional composition, with the MLA and Karnataka samples affected probably by higher mortality levels due to their specific characteristics. The second hypothesis relates presupposes that Kerala can be used as the prototype of India’s age and sex patterns of Covid-19 mortality. We can then adjust the intensity of mortality according to a Covid-19 mortality model based on previous studies on the age gradient of Covid-19 mortality across the word. The age factor is treated in greater detail in Appendix 2, but we will briefly introduce the readers to the main argument of the reasoning.

Covid-19 mortality by sex and age has now been measured in most countries in the world with disaggregated statistics published by national statistical bureaus.^**22**^ Beyond the overall observation that Covid-19 mortality rises with age and is more pronounced among males, the comparison of these mortality curves has highlighted their extreme regularity. Covid-19 mortality above age 20 follows everywhere the so-called Gompertz law of mortality (in which death rates can be expressed as an exponential function of age). ^**23 24 25**^ As a result of this strong mathematical regularity, death rates unknown at one given age can be extrapolated from death rates observed at other ages. Another advantage of the Gompertz modeling is its partition of mortality into two main components: its age gradient (i.e., the slope, or the *tempo* of mortality) that corresponds to variations by age, and its intensity (the level or *quantum* of mortality) that captures the overall severity at all ages. By adjusting the quantum of mortality, we can simulate the effects of higher or lower Covid-19 intensity at all ages while keeping age patterns (the tempo) fixed.

The age profile of Kerala’s Covid-19 death rates is perfectly in line with mortality patterns observed elsewhere, including in South Asia. We can use them and then adjust the quantum of mortality to deaths observed in other samples. The flexibility of the model enables us finally to estimate death rates in age groups that are not part of the original populations of the samples used, such as children or the elderly (see Appendix 2).

The different steps in the estimation procedure can now be described. We first compute age and sex Covid-19 death rates based on the Kerala sample and then derive from these rates a preliminary Gompertz model of Covid-19 mortality. This model is applied to the samples’ populations classified by sex and age and then adjusted to fit the number of deaths observed in these samples. The resulting adjusted mortality model is finally applied to India’s entire population by age and sex, yielding the projected national number of Covid-19 deaths. We further derive various comparative indicators of Covid-19 mortality and use for this purpose the demographic and Covid-19 data for Brazil, the USA and the world. The workflow is summarized in Figure 1, with further explanations and illustrations found in appendices 2 and 3.

**Figure 1:**
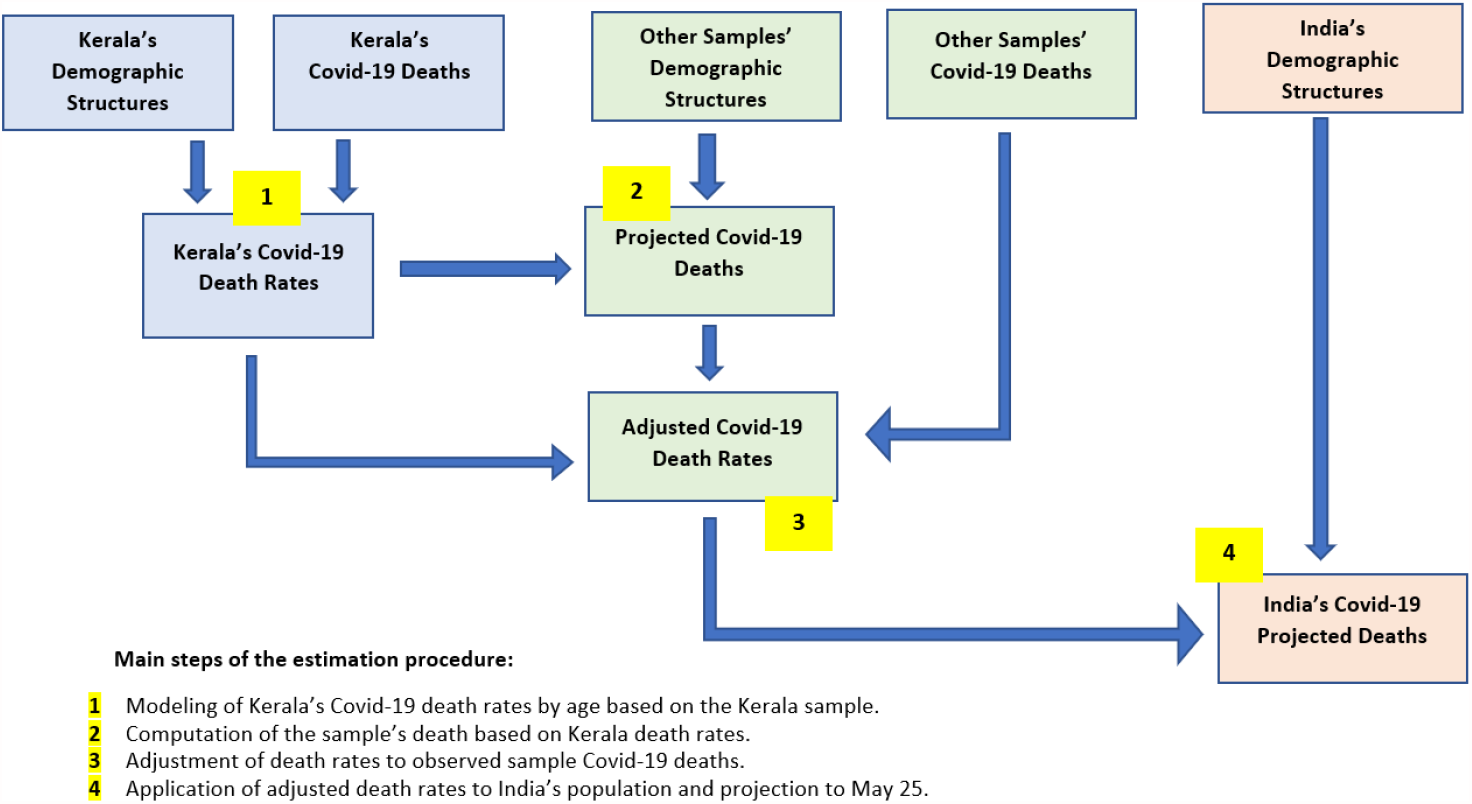
Estimation procedure.

## 4. Findings

Table 1 summarizes the results of the estimation procedures applies to the IR and MLA samples. The estimated total number of Covid-19 deaths in India till May 25, 2021, varies from 2.16 million (IR sample) to 2.45 million (MLA sample).

**Table 1:**
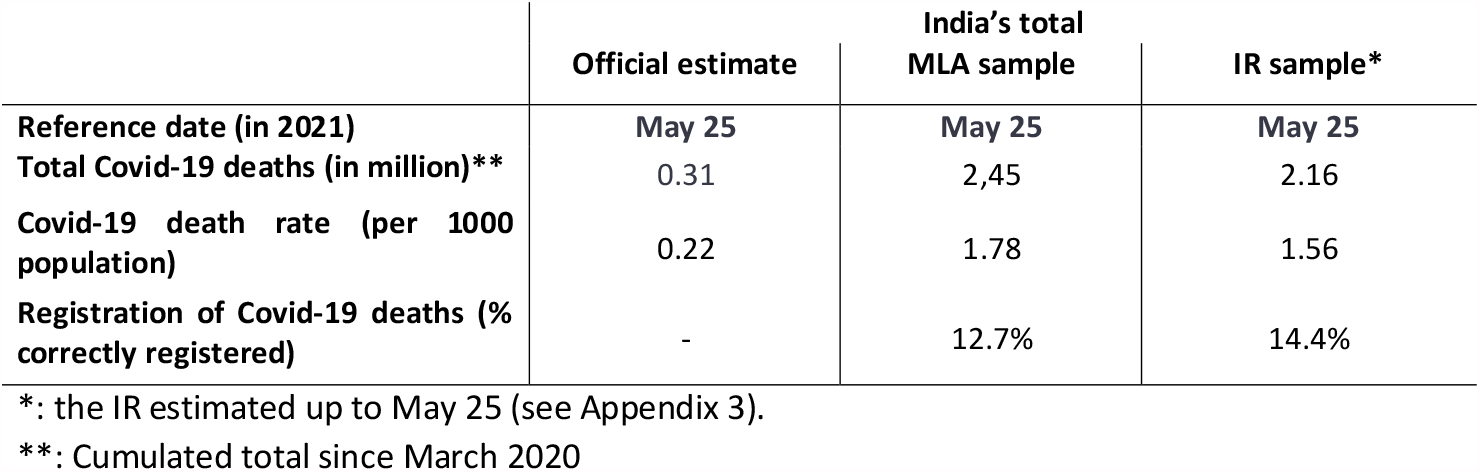
Estimates of Covid-19 death rates in India based on the IR and MLA samples.

|  | Official estimate | India's total<br>MLA sample | IR sample* |
| --- | --- | --- | --- |
| Reference date (in 2021) | May 25 | May 25 | May 25 |
| Total Covid-19 deaths (in million)** | 0.31 | 2,45 | 2.16 |
| Covid-19 death rate (per 1000 population) | 0.22 | 1.78 | 1.56 |
| Registration of Covid-19 deaths (% correctly registered) | - | 12.7% | 14.4% |

\*: the IR estimated up to May 25 (see Appendix 3).
\*\* : Cumulated total since March 2020

These figures correspond to a crude Covid-19 death rate ranging from 1.6 per 1000 (IR sample) to 1.8 (MLA sample). The overall mortality tally is 7 to 8 times as high as the official number of Covid-19 deaths.

We first note the global agreement of both estimates that put the death tally far above the official figures, at levels above million deaths. In addition, these estimates broadly to previous estimates mentioned earlier that put total Covid-19 mortality in India at levels close to 1.5-2 million deaths. Our estimates concur in confirming the serious doubts leveled at the reliability and exhaustivity of official Covid-19 death counts published so far and the suspicion of large-scale under-registration.

Absence of sample data by period prevents us, however, from computing the registration rate during the first and the second waves.

There are, however, variations in the estimates. The MLA sample yields results that are 14% higher than with the IR sample. This difference may reflect measurement issues, but above all the composition of each sample. As argued in Appendix 1, higher mortality observed among elected representatives (MLA sample) should be primarily attributed to their extreme exposure to contamination since the outbreak of the pandemic due to their continuous interaction with their electorate and the busy election schedule of 2020-21. Their elite status may not have offered politicians protection from the coronavirus and its deadly consequences. We should therefore regard the mortality level derived from the MLA sample as the upper bound of India’s Covid-19 mortality, applicable to India’s most exposed groups of frontline workers.

Comparatively, the mortality estimate based on the IR staff appears to capture more effectively the impact of the pandemic on India’s population. The social, economic and social diversity of this subpopulation reflects relatively well India’s heterogeneous population. The fact that part of the IR workforce was made of frontline workers also mirrors the large number of Indian workers directly exposed to the virus during the pandemic. The total of 2.16 million deaths drawn from the IR sample appears the best estimate yet of the overall impact of Covid-19 mortality in India from March 2020 until May 25, 2021.

The IR estimate gets a first confirmation from the Karnataka sample. This sample recorded 268 Covid-19 deaths among government schoolteachers in the Southern state of Karnataka. The application of the IR mortality patterns to the age and sex structure of Karnataka’s schoolteachers yield a preliminary total of 162 deaths. When we correct the latter figure for Karnataka’s relatively higher Covid-19 death rate (66% above India’s as per official estimates), we obtain a final estimate of 269 deaths among schoolteachers based on the IR mortality level. This figure is remarkably close to the observed number among Karnataka’s schoolteachers and reinforces the consistence of the figures based on the IR sample.

## 5. Discussion

All over the world, the identification and proper registration of Covid-19 deaths have been a challenge. Due to data restriction or severe underestimation in many countries, Covid-19 mortality has most probably also been severely underestimated in Brazil, Iran, Peru, Russia, or Turkey.^**26 27 28 29 30**^. In India’s case, the estimated mortality level is considerably higher than that reflected by the official statistics, by a factor of one to 7. This pronounced undercount is the joint product of the under-registration of adult deaths in India, the reluctance or inability of families or of local authorities to correctly identify Covid-19 deaths, and the faulty attribution of Covid-19 deaths to other causes. While this confirms the reservations about the quality of Covid-19 data, our estimate is higher than many figures previously circulated in the media.^**31**^ The salience of these estimates appears more clearly when set against figures from other countries (Table 2). With 2.2 million Covid-19 deaths by May 25, India now emerges as the country with the largest number of deaths in the world, well ahead of the USA, Brazil or Mexico. India’s Covid-19 death count would amount to 40 percent of the revised death total. Deaths have peaked around May 20 and have been declining in June 2021 and we can project the cumulative number of deaths in India until the end of the second wave. If an equivalent number of people succumbs during the rise and during the decline of the second wave–as was observed for the first wave in 2020–, the total death toll should be close to 3 million by August 2020.

**Table 2:**
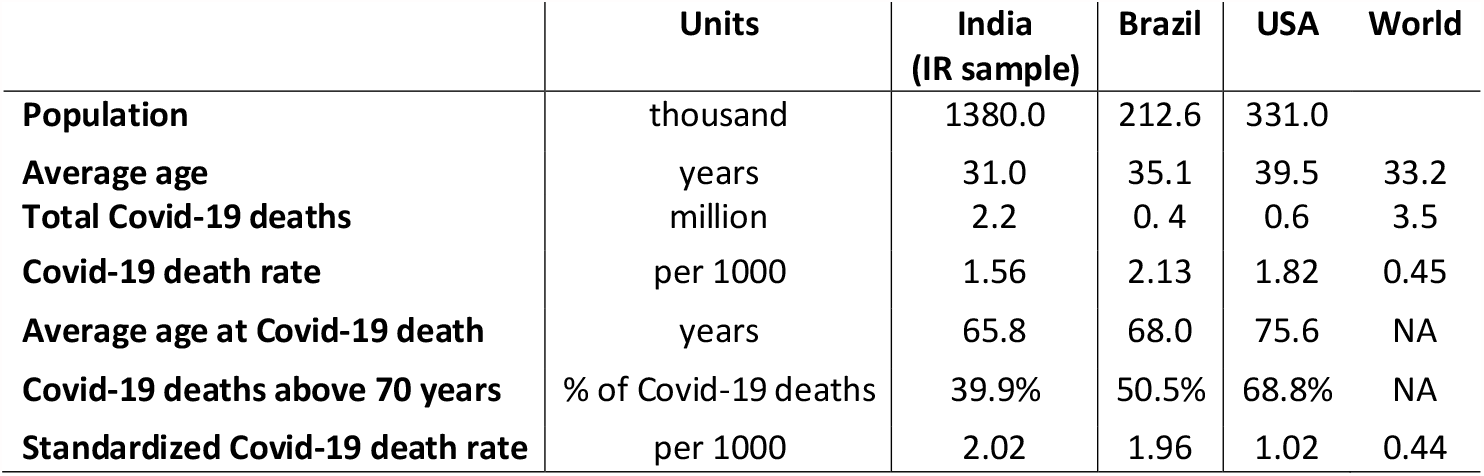
Comparative indicators of Covid-19 mortality.

Another important result of our estimation relates to Covid-19 mortality rates. The highest crude Covid-19 death rates per inhabitant are observed in Central Europe (Hungary, Czechia, Bosnia and Herzegovina), with rates ranging from 2.8 to 3.1 deaths per 1000. Belgium (2.1 per 1000) tops the Western European countries, with the USA (1.8 per 1000) and Brazil (2.1 per 1000) record the highest rates in North and South America, respectively. India’s revised Covid-19 death rate based on the IR sample stands now at 1.56 per 1000—a level well above the world average and close to that of badly hit countries in Western Europe or Latin America such as Argentina and France.

However, these crude Covid-19 death rates are directly affected by the age and sex composition of each population. Countries such as the USA with a larger proportion of population aged 60+, most vulnerable to Covid-19 mortality, are bound to register more deaths than relatively younger countries like Brazil or India *ceteris paribus*.^**32**^ This gets reflected in the average age at death of Covid-19 patients and the share of deaths above 70 years shown in Table 2. To offset the effect of demographic structures, we also show standardized Covid-19 death rates based on the world’s population in 2020 (see Appendix 2). These standardized death rates are not any longer affected by each country’s specific population size and its sex and age structure. India’s standardized Covid-19 mortality rate now stands at 2.02 per 1000, a level almost double that of the USA (1.02) and of European countries.

Brazil’s standardized Covid-19 death rate at 1.96 per 1000 is now below India’s. After age and sex standardization, India’s Covid-19 death rate is clearly among the highest in the world. The lack of proper statistics has therefore concealed the extreme severity of India’s Covid-19 pandemic thanks to the dramatic underestimation of its Covid-19 deaths and its relatively favorable age structure. In addition, India’s Covid-19 death toll is likely to continue rising till the end of the second wave.

The true extent of the outbreak of the coronavirus in India requires immediate attention and support from the international community to help Indian authorities respond to all aspects of this health crisis by combining non-pharmaceutical interventions and scale-up of vaccination to make them accessible to all, with an improved surveillance system to monitor the evolution of the pandemic.

As a final observation, we should stress the limitations of our approach. The estimates presented here remain intricately linked to the quality of information in the population samples. The IR sample is used here as the “best one yet”, but it is not the final sample. In the future, large new samples should ideally include subpopulations underrepresented here such as womenfolk, urban migrants, the peasantry and informal workers, and the slum population. They may also allow to better document the poorly understood socioeconomic and gender differentials in Covid-19 mortality. In addition, many questions remain unanswered such as the breakdown of Covid-19 deaths by sex or by region, the specific impact of the second wave and of the new Delta variant originating from India, and the contribution of local comorbidity pattern to the death toll.

## Data Availability

data available online

## Acknowledgements

a prior version of this paper benefited from comments from V. Balaji, N. Belorgey, G. Duthé, P. Heuveline, M. John, and F. Roubaud,

## Appendix 1: Sample characteristics

There are now many sources providing the number of Covid-19 deaths for specific communities, organizations, or localities. The idea is to identify reliable samples of deaths that may provide an alternative estimation of Covid-19 mortality risks. We have examined for that purpose most available samples of mortality statistics related to COVID-19 and checked them for reliability, exhaustivity and representativeness. The selection criteria are the following:

1. Samples based on well-defined populations (denominator).
2. Reliable estimation of the Covid-19 deaths (numerator)
3. Samples large enough for mortality estimates (robustness).
4. Populations with known age and sex structures (demographic composition)
5. Samples covering the entire pandemic since March 2020 (timescale).
6. Samples representative at all-India level (coverage).

We need samples that are large enough to provide robust estimates and refer to well-defined populations as we have basic demographic information, i.e. the age and sex distribution. We have unfortunately no specific sample so far on specific sections of the population such as farmers, factory workers, or the police. After examination, we discarded most sources that provided incomplete or potentially unrepresentative information. The following samples were rejected:

- **Indian Armed Forces** (119 deaths). It is a large sample, composed mostly of young men below 40, but we cannot use it for lack of detailed information on its demographic composition.
- **Medical community** (244 deaths among doctors during the second wave). It is a small sample, but it was rejected chiefly due to its extreme exposure to contamination during the pandemic. The same is true for other samples of health personnel.
- **Schoolteachers in Uttar Pradesh** (1621 deaths). We discarded this sample for several reasons: lack of demographic characteristics of the population at risk, lack of data for 2020, and suspiciously high mortality rate in 2021.
- **Bank employees of India** (1,300 deaths). This sample is not usable to the lack basic information on this sample’s demographic characteristics and may have been especially exposed to the pandemic.
- **Employees of Hindustan Aeronautics Limited** (100 deaths) and **civil aircraft pilots** (17 deaths). These samples are too small and undocumented.
- **Death claims from insurance companies** (35,500 deaths). This sample is incomplete in terms of deaths, and we don’t have the necessary information on its demographic composition. In addition, the insured population may not be representative of ordinary Indians.

We finally retained the following four samples which met our criteria in terms of reliability of death estimation, regional representativeness, and demographic characteristics.

1. **Kerala sample**: Dataset of deaths by age and sex in Kerala
2. **MLA sample**: Deaths of elected representatives
3. **IR sample**: Deaths of Indian railways personnel
4. **Karnataka sample**: Deaths of schoolteachers in Karnataka

In the following paragraphs, we describe the main characteristics of these samples and discuss their representativeness for estimating India’s Covid-19 mortality.

A. The **Kerala sample** is a dataset of 6339 Covid-19 deaths classified by age and sex. It has been compiled by the regional bureau of statistics and is available online as a database of individual deaths with relevant characteristics such as the date and place of occurrence, and the age and sex of the deceased.^**33**^ This estimate of Covid-19 deaths contradicts the apparent lack of excess mortality in 2020 derived from a straightforward examination of death statistics. This further highlights the risk of using excess mortality as an indirect indicator of the Covid-19 death toll.

This data source is unique for India. To the best of our knowledge, no other statistical office at regional or national level has ever published in the country disaggregated information on Covid-19 deaths in an accessible format. Moreover, the dataset is regularly updated by the addition of more recent deaths. Kerala is otherwise known for the quality of its demographic information, a feature related to its overall educational level. We use here data downloaded on May 18. As indicated by Figure A1, this dataset still reflects some amount of age heaping for ages ending with 0 and 5. We smoothed the individual age data for the population above 30 with a moving average over five successive years.

**Figure A1:**
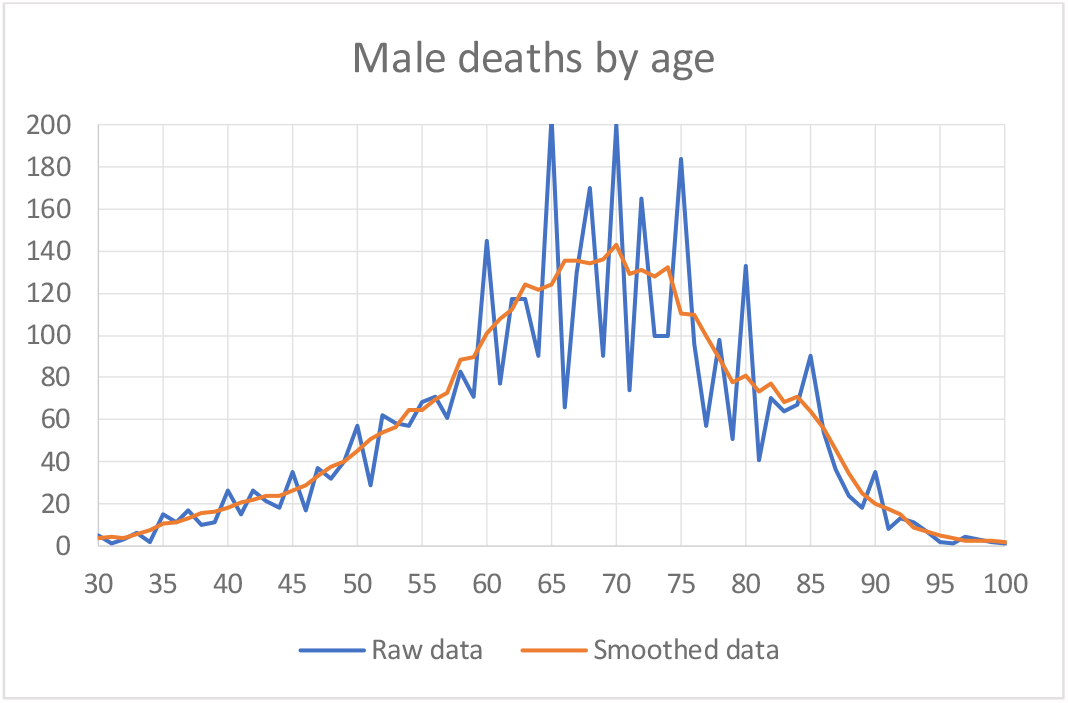
Distribution of male Covid-19 deaths by age, Kerala, May 18.

Kerala’s death reflects the strong age gradient of Covid-19 mortality. Deaths below 30 accounts only for 1.3% of all deaths. The number of deaths peaks at age 70, but the distribution of deaths among the elderly is directly affected by the original age distribution of Kerala.

We now use this series to compute the age- and sex-specific Covid-19 mortality rates by computing the ratio of death per person. In the absence of updated series, we use for this purpose the age and sex distribution of Kerala as estimated during the last 2011 census. The death rates for men and women are shown in Figure A2.

**Figure A2:**
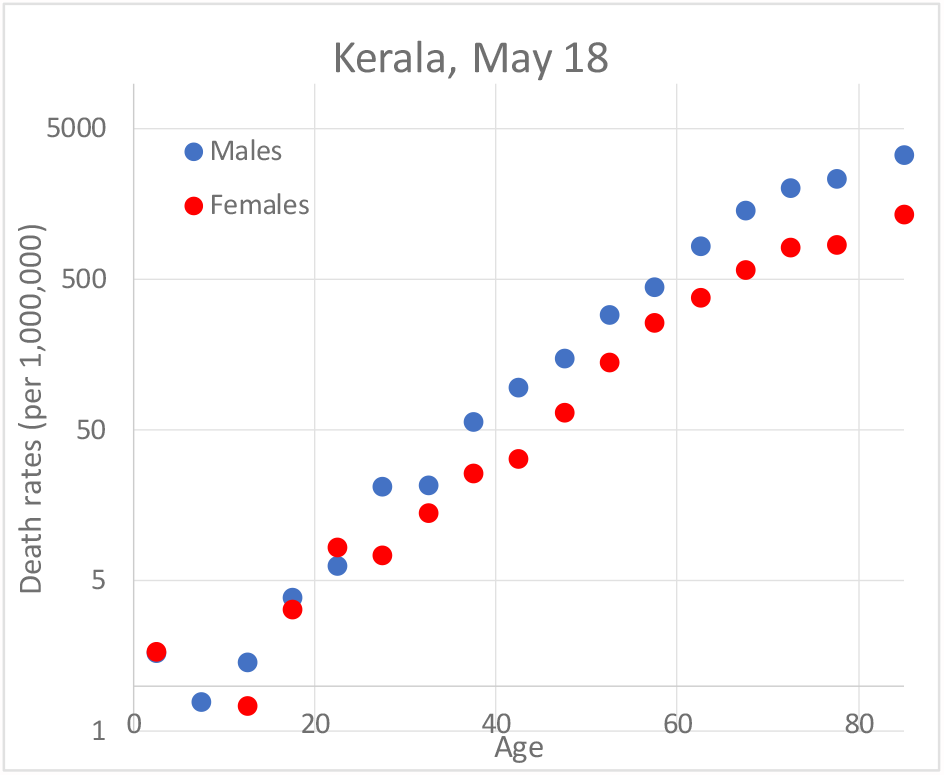
Covid-19 death rates by five-year age and sex, Kerala, May 18.

As we can see, the series appear relatively erratic for ages below age 15 due to the small number of reported deaths. There are for instance no reported Covid-19 deaths among girls aged 5-9 years. This is of limited importance since death rates at younger ages are almost negligible and do not influence in any way the overall Covid-19 death toll. It should also be stressed that since we have no verifiable information on the real Covid-19 under-registration in Kerala, this series cannot be used to summarize India’s overall mortality level.

These rates show, however, that Kerala’s mortality patterns follow the usual profile of Covid-19 mortality, with death rates higher among men and fast rising with age (note the log scale used here). We will use these features further below to model the shape of Covid-19 mortality in India (see Appendix 2).

The **MLA sample** is relatively small sample of 5,837 individuals. Individuals in this sample are the elected representatives of India, starting with the MLAs (Members of the Legislative Assembly) sitting in the regional *Vidhan Sabha*s. We have added the national parliament, i.e. the *Lok Sabha* (534 members) and the *Rajya Sabha* (226 members). MLAs represent 85 % of the entire sample. About 0.5% representatives are missing from the sample due to lack of information (or vacant seats). Elected representatives cover the entire territory of India, even if 5 smaller territories with no legislative assemblies.

The age and sex composition is available from published databases such as Trivedi Centre of Ashoka University and from the websites of the Lok Sabha and the Rajya Sabha.^**34 35 36**^ The overall age distribution is applied to the part of the sample (20%) for which age information is missing. Women account for only 7.1% of representatives. The mean ages are relatively high at 56.2 years for men and 50.4 for women. This makes them a relatively vulnerable group in view of its advanced age, almost twice higher than the entire population of India.

Deaths by Covid-19 of elected representatives have been systematically covered by the national or regional press, precise information on the date and location of the event. Deaths of elected representatives also lead to vacancies or to by-elections reported in the media. We used a list of Covid-19 deaths in India to crosscheck our results. A total of 43 deaths by Covid-19 of elected representatives were reported from 2020 till 25 May 2021, which points to a rather high fatality rate of 74 per 1000 in this subpopulation. Incidentally, a simple comparison shows this overall death rate to be comparable to Covid-19 death rates for the American population aged 55-64 years (Figure A4).

The average age at death in the MLA is 65.6 years, quite higher than the estimated figure for India (Table 2). 60 % of these deaths occurred during the first wave (i.e. before February 2021) as against 57% according to the official estimates of Covid-19 deaths for India. Figure A3 shows that the cumulative distribution of deaths in the MLA sample mirrors that of officially reported Covid-19 deaths in India.

**Figure A3:**
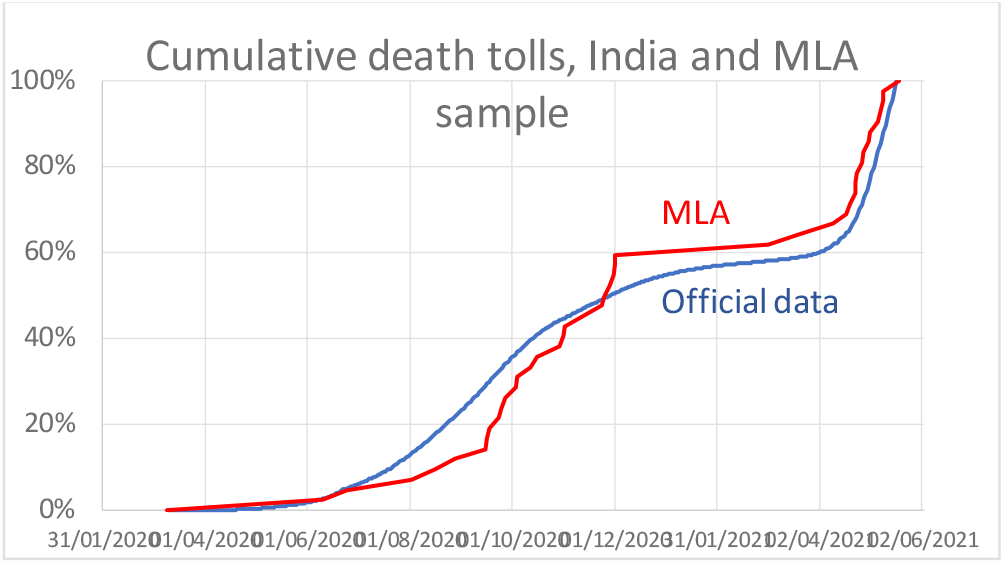
Cumulative Covid-19 deaths from February 2020 to 20 May 2020, Indian official estimates and MLA sample percentage of total deaths).

This is an admittedly small sample, but of high quality and precision. The MLA sample corresponds to a high stratum of India society and represents one of India’s most visible elite groups. They are primarily city-based and apart from their relatively high salaries, they enjoy numerous added benefits such as housing, home staff, specific allowances, traveling and energy reimbursement, access to medical facilities, and pension benefits. This situation was reflected notably by the fact that a vast majority of Covid-19-related deaths among MLAs took place in the most reputed Indian hospitals across the country.

Yet, this population most probably was subjected to a very level of exposure to contamination with the population of their constituencies. Many politicians initially expressed support for unproven local remedies and did not follow standard protective measures based on social distancing. Like politicians elsewhere in the world, they continued to meet hundreds of people on a regular basis in meetings of various sizes and many got contaminated early on during the pandemic. But the activities of regional and national representatives in India never stopped throughout the pandemic, especially during the different elections rounds in November 2020 and April 2021 in five states and territories and the local (*panchayat*) elections held in India’s largest state in April 2021 during the middle of the second wave. These elections (and a major pilgrimage held at the same period) were blamed for the accelerated diffusion of the coronavirus in North India during the second wave of 2021. In spite of their easier access to quality medical care, the human cost of the pandemic on political leaders appears to have been considerable. We therefore regard the mortality level derived from the MLA sample as the upper bound, applicable mostly to India’s most Covid-19-vulnerable groups.

C. The **IR sample** (where IR stands for Indian Railways) is a large sample that relates to the employees of the Indian Railways. With 1.3 million employees in 2020, the Indian Railways is in fact one of the world’s biggest employers.^**37**^ The IR network covers the entire territory of the country, except for insular and mountainous regions s. It announced on May 10 that 1,952 staff died of Covid-19 (1.5 per 1000), a figure that should refer to the previous Friday, May 7 at the latest. For comparative purposes, the total will be projected until May 25 in proportion to the rise of official Covid-19 deaths.

Covid-19 deaths are likely to be adequately numbered for two main reasons. First, the IR is an organization with a unique internal health system (the Indian Railway Health Service with 13,600 hospital beds, 2,600 doctors and 41,000 other health staff), though which employees and their family enjoy access to quality healthcare, including in time of Covid-19 crisis. Second, the main union of the Indian Railways has long been calling for a Covid-19 higher compensation for deceased employees, a move that has probably encouraged families to adequately report cases of Covid-19 deaths.

As most Indian organization, the IR does not publish statistics on the demographic composition of its workforce. The only information included in its regular yearbooks is the share of female employees, standing at 99 thousand workers in 2020 (7.9%). There are, however, enough information to reconstruct the age structures of its workforce, viz. the average age of employees (45 years) and the share of employees aged 50 or above (40%).^**38 39**^ Using these two parameters, we have fitted a linear distribution by five-age groups from ages 20 to 60 (ages for recruitment and retirement). Less regular age distributions with similar demographic characteristics have been tested with no significant bearing on mortality estimation.

The IR sample represents a share of the India’s workforce that is quite heterogeneous and spread across regions. It includes all socioeconomic groups (from peons to engineers) and covers all social groups, including lower-status groups (Other Backward Classes, Dalits or Tribals) who enjoy job reservations. However, this sample does not cover workers from the informal economy.

Overall, this group of IR workers may be slightly more privileged than the rest of the population for various reasons: it is a more urban population (following the railway grid) made of permanent employees (public servants) that have access notably to income security, housing allotment or subsidies, dedicated healthcare system, provident fund and pension benefits. Yet, it is also a population that may have been exposed to higher risks of Covid-19 contamination due to the volume of passenger traffic (8.1 billion in 2019-20). As proof, the death rate is twice as high among station masters (2.9 per 1000) as among the rest of the IR staff (1.5 per 1000). Yet, most of the IR staff are not frontline workers, but engineers, clerks, mechanics, pilots, and other technicians (such as *pointsmen* or *gangmen)* that are not in contact with the public. The IR traffic has also been repeatedly interrupted during the pandemic, most notably during the initial lockdown in March-May 2020 during which almost no train ran through India. For all these reasons, linked both to its social and regional composition and various exposure levels to contamination, this subpopulation is the closest we found to the Indian public and will provide what we consider our best estimates yet of Covid-19 mortality in India.

D. The **Karnataka sample** is a sample of 268 deaths attributed to Covid-19 among schoolteachers of southern state of Karnataka. This number of deaths was reported in the press on May 13. Albeit small, this number can be related to the demographic structure of the Karnataka’s 196,163 teachers. The sex and school-level composition of teachers in Karnataka can be found in the most recent administrative report.^**40**^ Unlike the MLA and IR samples, a majority of them are women (52.6%) In addition, a special study provides an estimate of the average age of the schoolteachers (39.5 years).^**41**^ This number closely corresponds to the official age limit of the profession (from 21 to 59 years). While teachers may represent vulnerable frontline workers, schools in Karnataka were closed for most of the pandemic’s period and they may not have been more exposed to the virus than the general population.

Karnataka apparently recorded a larger proportion of cases and deaths than the rest of India, with the metropolitan region of Bengaluru severely hit by both waves of the pandemic. For this reason, this sample cannot be used to estimate the national mortality level. However, it can be used to test the validity of mortality estimates derived from the IR sample.

## Appendix 2: The Age and Sex Factor

Existing Covid-19 mortality statistics in India are limited to cumulative totals classified by administrative division. Covid-19 cases are tabulated elsewhere by age group and sex and statistical offices even make these numbers available as datasets of individual deaths as is common in Latin American countries.^**42**^ Such disaggregated data have allowed for the computation and analysis of Covid-19 death rates by age and sex in various countries.^**43**^ This led to recognition of the strong age gradient of Covid-19 mortality observed throughout the world. Research on Covid-19 mortality has shown that the progression of mortality with age is very rapid, faster than for ordinary mortality. For instance, there is a tenfold increase in Covid-19 mortality risk when age increases by 20 years. This means that Covid-19 mortality risk is ten times higher at age *a+10* than at age *a*. The age composition of the population at risk plays therefore a crucial role in the overall death toll, a feature already illustrated by the record Covid-19 death rates observed in North America and European countries.

The regularity of mortality patterns has also highlighted that Covid-19 mortality follows the typical features of the so-called Gompertz law.^**44**^ In this classical mortality model, age-specific mortality rates can be approximated as an exponential function of age. The standard Gompertz equation is:

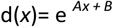

where d(*x*) is the death rate at age *x*, while A and B are parameters.

Coefficient A (>0) captures the age gradient of mortality, i.e. mathematical slope (the *tempo* of mortality). On the contrary, coefficient B corresponds to the overall intensity (the *quantum* of mortality) and affects mortality levels at all age. These two coefficients allow for a complete parameterization of age-specific mortality rates according to their tempo (early vs. late mortality) and quantum (low vs. high mortality). This type of modeling is therefore applicable to Covid-19 mortality rates by age.

**Figure A4:**
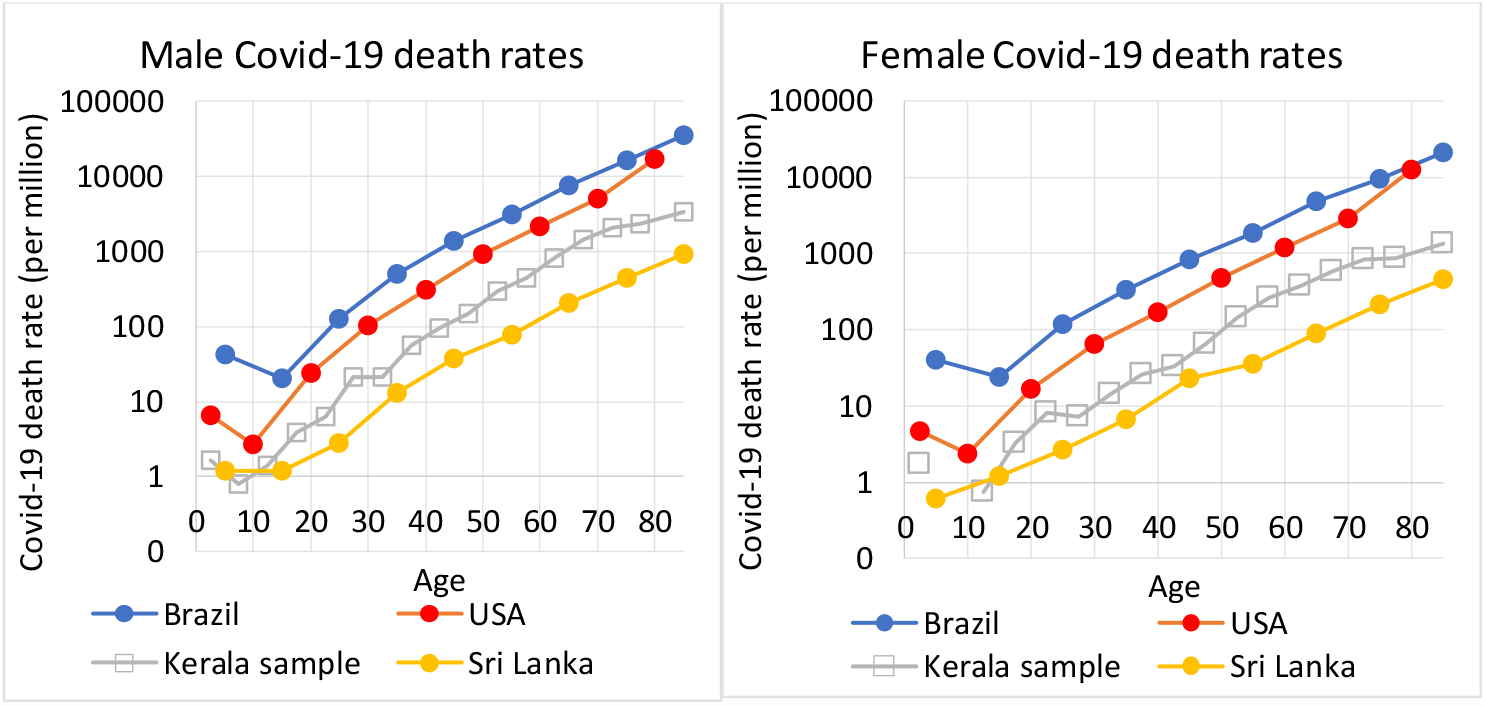
Covid-19 death rates by age and sex, Brazil, Kerala, Sri Lanka and USA, May 2020.

Figure A4 illustrates the common Covid-19 mortality profiles across the world. We have computed the age-specific Covid-19 death rates based on available cumulative data of deaths by age for Brazil and the USA, the countries with the largest number of Covid-19 casualties in the world.^**45 46**^ Like India, Bangladesh, Nepal, and Pakistan have failed to publish the series of Covid-19 deaths disaggregated by age and sex. The only South Asian exception is Sri Lanka, where the Epidemiology Unit of the Ministry of Health releases every week an updated count of deaths by age and sex.^**47**^

The data refer to mid-May 2021 and follow the age format specific to each country. The death rates are computed based on the United Nations population estimates of Brazil, Sri Lanka, and the USA for 2020. We see strong exponential relation between age and Covid-19 death rates in each country, but we also notice the somewhat parallel profile of all mortality patterns. Kerala’s profile stands halfway between Sri Lanka’s low Covid-19 death rates and the highest levels recorded in the USA and Brazil.

Further analysis also shows that the progression of Covid-19 death rates tends to decelerate above 70.^**48**^ This feature is noticeable for Brazil, Kerala and among Sri Lankan males, but not for the USA. Modeling for mortality among children mortality also tends to be difficult. Not only are deaths at younger ages very few, but there seems to a slight Covid-19 mortality surge below age 5. This turnaround mirrors the cusp of general mortality which is invariably higher at ages 0-4 than at ages 5-9.

The previous figures showed that death rates computed on the Kerala sample follow the expected Gompertz profile and are remarkably like patterns is found elsewhere, notably the slope (tempo) of mortality. In order to model more precisely Kerala Covid-19 mortality at all ages, we introduce here a slightly more complex Gompertz equation with four parameters:

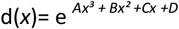

where d(*x*) is the death rate at age *x* and *A, B and C* parameters capturing the non-linear age gradient (tempo) of Covid-19 mortality. *D* represents the overall quantum of Covid-19 mortality. This polynomial parameterization provides a better adjustment to both the turnaround of Covid-19 mortality at young ages and its deceleration at older ages (Figure A5).

**Figure A5:**
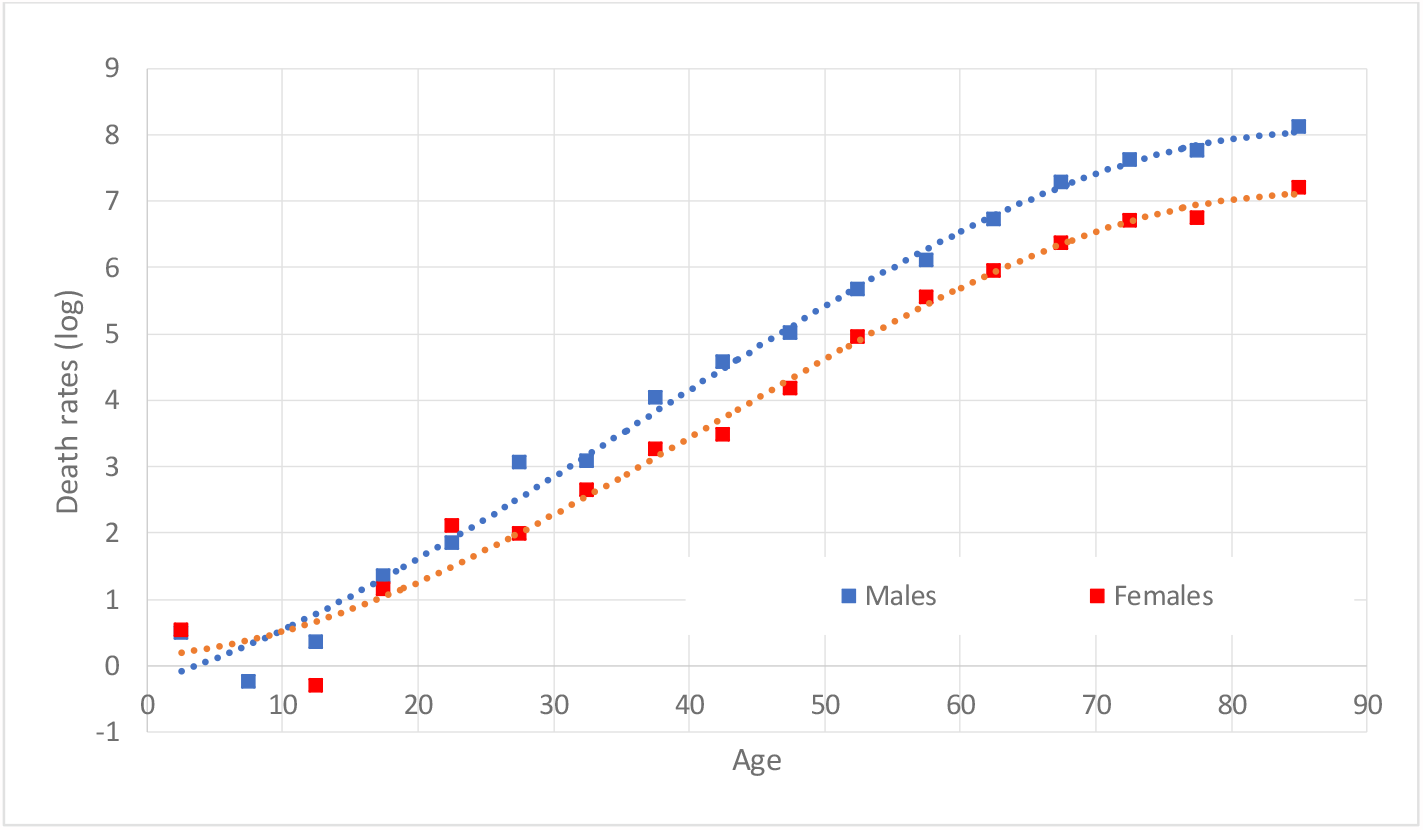
Observed and modeled Covid-19 death rates by age and sex, Kerala, May 2020.

This model provides an almost perfect fit for adult ages, with r^2^= .995 (males) and .992 (females) for ages above 15 years. It is less precise for children because of the near absence of cases of child deaths and the cusp observed at ages 5-9 years. The overall correlation remains extremely strong with r^2^ at .990 for male rates and .981 for female rates. The gap for death rates below age 15 has almost no repercussion on the overall mortality toll due to the extremely low level of mortality risk at young ages (death rates < 3 per million for the population aged less than 15).

The advantages of this generic model of Covid-19 mortality by age and sex for our estimation procedures are twofold:

- The model allows for the estimation of Covid-19 death rates at all ages–age bands not included in our samples–thanks to the coefficients *A, B* and *C*.
- The model can be adjusted to fit different overall mortality levels with the help of coefficient *D*.

## Appendix 3: Computations

In this section, we illustrate the estimation procedures with the case of the IR sample.

We first distribute the sample population by sex and age according to available information. In the case of the IR sample, we use four types of information: the population size, the share of women, the average age of the workforce and the proportion among them aged 50+. To conform to general age regulations pertaining to recruitment and retirement in the Indian railways, the sample population is strictly contained in the 20-59 age band. We use here a simple linear distribution by age, but other imputations do not affect the results. The imputed population distribution is shown in columns 2-3 of Table 1.

We then apply mortality rates by age and sex derived based on the Kerala model (see Appendix 2) shown in columns 4-5 of Table 1, with the number of projected deaths shown below. The death by age and sex is then adjusted upward (columns 6-7) to match the observed death rates reported by the Indian Railways on May 10 (i.e., a total of 1,952 deaths).

We finally applied the revised model extended to all ages from 0-4 to 80+ to the entire age and sex structure of India. India’s figures shown in columns 8-9 are drawn from the United Nations estimates published in 2019 (World Population Prospects). This simulation yields a total number of 1.65 million deaths up to May 7 (columns 10-11). For comparative purposes, this total will be projected to 2.16 million on May 25 using the trend observed of official Covid-19 deaths during this period (+30.7%).

We finally use the world age and sex structure also derived from the United Nations estimates to standardize the Covid-19 death rate. To do that, we recompute each sex and age group by applying the proportion observed in the world in 2020 to India’s population total (columns 12-13). We then apply the revised mortality model (columns 6-7) to India’s standardized age and sex distribution to get the standardized deaths (columns 14-15), yielding the standardized Covid-19 death rate.

The other computations derived for the other MLA and Karnataka samples follow an identical pattern.

**Table A.1:** Estimation procedure of Covid-19 mortality in India based on the IR sample (May 7, 2020).

|  | Indian Railways workforce |  | Kerala Covid-19 death rates |  | Adjusted death rates |  | India's population |  | Projected Covid-19 deaths |  | India's standardized population |  | Standardized Covid-19 deaths |  |
| --- | --- | --- | --- | --- | --- | --- | --- | --- | --- | --- | --- | --- | --- | --- |
|  | Male | Female | Male | Female | Male | Female | Male | Female | Male | Female | Male | Female | Male | Female |
| Unit | persons |  | per million |  | per million |  | thousand |  | deaths |  | thousand |  | deaths |  |
| Source | IR sample |  | Kerala sample |  |  |  | United Nations |  |  |  |  |  |  |  |
| 1 | 2 | 3 | 4 | 5 | 6 | 7 | 8 | 9 | 10 | 11 | 12 | 13 | 14 | 15 |
| Age group |  |  |  |  |  |  |  |  |  |  |  |  |  |  |
| **0-4** |  |  |  |  | *7* | *9* | 61,228 | 55,651 | 429 | 521 | 61,864 | 58,160 | 434 | 544 |
| **5-9** |  |  |  |  | *10* | *11* | 61,877 | 56,105 | 647 | 628 | 60,713 | 56,921 | 634 | 637 |
| **10-14** |  |  |  |  | *17* | *15* | 66,303 | 59,853 | 1113 | 891 | 58,689 | 54,842 | 985 | 816 |
| **15-19** |  |  |  |  | *29* | *22* | 66,671 | 59,375 | 1912 | 1287 | 56,059 | 52,325 | 1608 | 1134 |
| **20-24** | 48,356 | 4156 | 6.7 | 4.5 | 51 | 34 | 64,866 | 57,639 | 3338 | 1960 | 54,580 | 51,183 | 2808 | 1740 |
| **25-29** | 75,781 | 6513 | 12.5 | 7.4 | 96 | 57 | 62,039 | 55,358 | 5936 | 3135 | 54,185 | 51,100 | 5185 | 2893 |
| **30-34** | 103,207 | 8870 | 23.9 | 12.9 | 182 | 99 | 58,875 | 53,301 | 10,718 | 5251 | 54,748 | 52,456 | 9966 | 5168 |
| **35-39** | 130,632 | 11,227 | 45.9 | 23.1 | 350 | 176 | 53,994 | 49,466 | 18,884 | 8721 | 48,943 | 47,513 | 17,117 | 8377 |
| **40-44** | 158,058 | 13,583 | 87.8 | 41.9 | 670 | 320 | 46,631 | 43,589 | 31,234 | 13,929 | 44,152 | 43,269 | 29,574 | 13,827 |
| **45-49** | 185,483 | 15,940 | 165.5 | 75.7 | 1262 | 578 | 40,757 | 38,684 | 51,442 | 22,340 | 42,708 | 42,160 | 53,905 | 24,347 |
| **50-54** | 212,909 | 18,297 | 302.9 | 134.4 | 2310 | 1025 | 35,203 | 33,673 | 81,318 | 34,498 | 39,411 | 39,509 | 91,039 | 40,477 |
| **55-59** | 240,334 | 20,654 | 531.5 | 230.4 | 4053 | 1757 | 30,150 | 29,106 | 122,195 | 51,126 | 34,030 | 34,635 | 137,922 | 60,839 |
| **60-64** |  |  |  |  | *6729* | *2866* | 24,695 | 24,196 | 166,174 | 69,339 | 27,827 | 29,205 | 187,253 | 83,693 |
| **65-69** |  |  |  |  | *10,436* | *4380* | 19,168 | 19,092 | 200,032 | 83,618 | 22,828 | 24,911 | 238,223 | 109,102 |
| **70-74** |  |  |  |  | *14,922* | *6174* | 11,628 | 12,464 | 173,510 | 76,950 | 15,436 | 17,968 | 230,326 | 110,932 |
| **75-79** |  |  |  |  | *19,418* | *7903* | 7,063 | 8,021 | 137,149 | 63,386 | 9,694 | 12,221 | 188,237 | 96,574 |
| **80+** |  |  |  |  | *23,481* | *9231* | 5,953 | 7,332 | 139,781 | 67,680 | 9,903 | 15,857 | 232,529 | 146,376 |
| **Population by sex** | 115,4760 | 99,240 |  |  |  |  | 717,101 | 662,905 |  |  | 695,770 | 684,236 |  |  |
| **Deaths by sex** |  |  | 247 | 9 | 1880 | 72 |  |  | 1,145,813 | 505,261 |  |  | 1,427,745 | 707,478 |
| **Total deaths** |  |  | **256** |  | **1,952** |  |  |  | **1,651,073** |  |  |  | **2,135,223** |  |
| **Covid-19 death rate (per 1000)** |  |  |  |  |  |  |  |  | Crude rate 1.20 |  |  |  | Standardized rate 1.55 |  |

Columns:
2-3: IR workforce age and sex distribution
4-5: Covid-19 death rates by age and sex derived from the Kerala sample.
6-7: Adjusted from preceding columns to fit the observed death among the IR sample (*rates in italics* used only for India's population )
8-9: Age and sex distribution of India's population in 2020 (United Nations estimates)
10-11: Projected Covid-19 deaths on May 7 computed from columns 6-7 and 8-9
12-13: Age and sex distribution of India's population standardized by the world's population (United Nations estimates)
14-15: Standardized Covid-19 deaths computed from columns 6-7 and 12-13

## References

1. India’s COVID-19 emergency. Lancet (London, England). 2021 May 8;397(10286):1683.

2. https://www.worldometers.info/coronavirus/#countries

3. https://www.reuters.com/world/india/indias-covid-crisis-pushes-up-cost-living-dying-2021-05-14/

4. Kumar GA, Dandona L, Dandona R. Completeness of death registration in the Civil Registration System, India (2005 to 2015). The Indian journal of medical research. 2019 June;149(6):740.

5. https://www.dw.com/en/india-coronavirus-death-toll/a-57338733

6. https://scroll.in/article/994619/india-is-undercounting-covid-19-deaths-heres-how-to-work-around-the-problem

7. https://science.thewire.in/health/india-mccd-comorbidities-covid-19-deaths-undercounting/

8. https://timesofindia.indiatimes.com/india/explained-why-indias-covid-19-data-is-vastly-undercounted/articleshow/82366707.cms

9. https://www.thehindu.com/opinion/op-ed/the-scale-of-gujarats-mortality-crisis/article34636681.ece

10. Bhattacharya P, Banerjee D, Rao TS. The “untold” side of COVID-19: Social stigma and its consequences in India. Indian journal of psychological medicine. 2020 Jul;42(4):382–6.

11. Joshi B, Swarnakar P. Staying away, staying alive: Exploring risk and stigma of COVID-19 in the context of beliefs, actors and hierarchies in India. Current Sociology. 2021 Feb 24:0011392121990023.

12. Islam A, Pakrashi D, Vlassopoulos M, Wang LC. Stigma and misconceptions in the time of the COVID-19 pandemic: A field experiment in India. Social Science & Medicine. 2021 Apr 28:113966.

13. Mukherji R. Covid vs. Democracy: India’s Illiberal Remedy. Journal of Democracy. 2020;31(4):91–105.

14. https://indianexpress.com/article/india/prabhat-jha-lack-of-death-data-prolongs-pandemic-survey-villages-7328846/

15. https://indianexpress.com/article/opinion/india-covid-death-toll-opinion-7348936/

16. Estimates by Murad Banaji: https://www.theindiaforum.in/article/estimating-covid-19-fatalities-india

17. Estimates by The Economist: https://www.economist.com/briefing/2021/05/15/there-have-been-7m-13m-excess-deaths-worldwide-during-the-pandemic

18. Estimates by the Institute for Health Metrics and Evaluation: http://www.healthdata.org/special-analysis/estimation-excess-mortality-due-covid-19-and-scalars-reported-covid-19-deaths

19. Estimates by the New York Times: https://www.nytimes.com/interactive/2021/05/25/world/asia/india-covid-death-estimates.html

20. Leffler, Christopher; Preliminary Analysis of Excess Mortality Suggests That Over 1.7 Million May Have Perished in Relation to the Covid-19 Pandemic in India: More Than in Any Other Country, preprint, June 2021 https://www.researchgate.net/publication/352170008

21. https://www.nytimes.com/2021/05/23/world/asia/coronavirus-kerala.html

22. Data compilation on Covid: https://population-europe.eu/network/news-network/ined-creates-website-international-data-demography-deaths-related-covid-19

23. Guilmoto CZ. COVID-19 death rates by age and sex and the resulting mortality vulnerability of countries and regions in the world. MedRxiv. Preprint, July 2020.

24. Ohnishi A, Namekawa Y, Fukui T. Universality in COVID-19 spread in view of the Gompertz function. Progress of Theoretical and Experimental Physics. 2020 Dec;2020(12):123J01.

25. Shapiro, V. COVID-19 Sex-Age Mortality Modeling - A Use Case of Risk-Based Vaccine Prioritization. 2021, https://doi.org/10.31235/osf.io/5c8bd

26. https://abcnews.go.com/International/data-suggests-russias-coroanvirus-deaths-higher-reported/story?id=70683286

27. Kisa S, Kisa A. Under‐reporting of COVID‐19 cases in Turkey. The International Journal of Health Planning and Management. 2020 Sep;35(5):1009–13.

28. Do Prado MF, de Paula Antunes BB, Bastos LD, Peres IT, Da Silva AD, Dantas LF, Baião FA, Maçaira P, Hamacher S, Bozza FA. Analysis of COVID-19 under-reporting in Brazil. Revista Brasileira de terapia intensiva. 2020 Apr;32(2):224.

29. https://www.reuters.com/world/americas/peru-almost-triples-official-covid-19-death-toll-after-review-180000-2021-05-31/

30. https://www.bbc.com/news/world-middle-east-53598965

31. https://www.nytimes.com/interactive/2021/05/25/world/asia/india-covid-death-estimates.html

32. Heuveline P, Tzen M. Beyond deaths per capita: comparative COVID-19 mortality indicators. BMJ open. 2021 Mar 1;11(3):e042934.

33. Kerala Covid Dashboard: https://dashboard.kerala.gov.in/deaths.php

34. Indian Elections Dataset, https://tcpd.ashoka.edu.in/lok-dhaba/

35. Lok Sabha website: https://loksabha.nic.in/

36. Rajya Sabha website: https://rajyasabha.nic.in/

37. Indian Railways, Year book 2019-20, Government of India, Rail Mantralaya, Ministry of Railways, 2020. https://indianrailways.gov.in/railwayboard/uploads/directorate/stat_econ/Annual-Reports-2019-2020/Year-Book-2019-20-English_Final_Web.pdf

38. Thirteenth Report Standing Committee on Railways (2016-17) (Sixteenth Lok Sabha) Ministry of Railways (Railway Board) Demands For Grants (2017-18) Presented to Lok Sabha on 10.03.2017, Laid in Rajya Sabha on 10.03.2017, Lok Sabha Secretariat New Delhi March, 2017.

39. Sindhu Bhattacharya “Record budget, but railways’ finances weighed down by big pension burden”, The Federal, February 2021. https://thefederal.com/budget-2021/record-budget-but-railways-finances-weighed-down-by-big-pension-burden/

40. Department of Primary and Secondary Education, School Education in Karnataka 2018-19, Bengaluru, 2020.

41. Kallan Gowda, Anjini Kocha, CS Nagabhushana and N. Raghunathan, Compensating Policies for Small Schools and Regional Schooling Inequalities: Class size and multi-grade teaching in India, Working paper n°455, Stanford University, Stanford, 2012.

42. Cepal N. Demographic Observatory Latin America and the Caribbean 2020: COVID-19 mortality. Evidence and scenarios., https://repositorio.cepal.org/handle/11362/46641

43. Bauer P, Brugger J, Koenig F, Posch M. An international comparison of age and sex dependency of COVID-19 Deaths in 2020-a descriptive analysis. medRxiv. 2021.

44. Guilmoto CZ. COVID-19 death rates by age and sex and the resulting mortality vulnerability of countries and regions in the world. MedRxiv. 2020.

45. Registro Civil do Brasil: https://transparencia.registrocivil.org.br/especial-covid

46. enter for Disease Control: https://data.cdc.gov/NCHS/Provisional-COVID-19-Death-Counts-by-Sex-Age-and-S/9bhg-hcku

47. Epidemiology Unit: http://www.epid.gov.lk/web/index.php?option=com_content&view=article&id=233&lang=en

48. Demombynes G. COVID-19 Age-Mortality Curves Are Flatter in Developing Countries, working paper, World Bank, 2020.

